# Epidemiological rather than macro-economic factors correlate with socioeconomic inequalities in HIV testing in 16 sub-Saharan African countries

**DOI:** 10.1101/2021.09.22.21263940

**Authors:** Pearl Anne Ante-Testard, Laura Temime, Kévin Jean

**Affiliations:** Laboratoire MESuRS, Conservatoire national des Arts et Métiers, Paris, France; Unité PACRI, Institut Pasteur, Conservatoire national des Arts et Métiers, Paris, France; MRC Centre for Global Infectious Disease Analysis, Department of Infectious Disease Epidemiology, Imperial College London, United Kingdom

**Keywords:** HIV, HIV testing, socioeconomic inequalities, contextual factors, sub-Saharan Africa, decomposing inequalities

## Abstract

In order to reach the first 95 (i.e., 95% of people living with HIV having knowledge of their status) of the 2030 UNAIDS 95-95-95 targets, it is crucial to better understand the contextual or structural factors driving socioeconomic inequalities in HIV testing uptake. It is still unclear whether they are mostly influenced by epidemiological or by macro-economic factors. Here, to shed light on this issue, we measured and decomposed socioeconomic inequalities in HIV testing in sub-Saharan Africa in relation to contextual factors using a novel method, the Recentered Influence Function decomposition method. Indeed, we found that HIV testing uptake was more concentrated among the rich in 12 of 16 sub-Saharan African countries based on population-based surveys. The level of the HIV epidemic seems to drive the level of response of HIV testing programs, rather than the per capita Gross Domestic Product of a country (i.e., national indicator of economic development). Our results suggest that when responding to the HIV epidemic, there is a need to monitor and assess inequalities in addition to monitoring HIV incidence and prevalence.

---

As the entry point to many HIV prevention and care services, HIV testing constitutes the first 95 (95% of people living with HIV will know their status) in the UNAIDS ambitious 95-95-95 targets by 2030 in ending the AIDS epidemic [1]. However, socioeconomic inequalities have been well documented in HIV testing, especially in sub-Saharan Africa (SSA), hindering the design of effective and efficient testing strategies. In order to increase testing uptake, better understanding the contextual drivers of these inequalities is necessary. For instance, it is unclear whether they are mostly influenced by epidemiological or by macro-economic factors. Here, to shed light on this issue, we measured and decomposed socioeconomic inequalities in HIV testing in SSA in relation to contextual factors.

We used data from the Demographic and Health Surveys conducted between 2008 and 2016 in a set of 16 SSA countries based on a previous study [2]. DHS are standardized nationally representative population-based surveys regularly conducted in low- and middle-income countries to collect data over a wide range of sociodemographic and health indicators including HIV and AIDS indicators. They have a multistage sampling design with household as sampling units. Individuals aged 15-59 years (majority 15-49 years) in selected households are generally eligible to be included in the survey. Country-specific per-capita Gross Domestic Products (GDP) of the corresponding survey years were obtained from the World Bank official website (https://www.worldbank.org/).

We calculated the country-specific Erreygers Concentration Index (ECI, values range from -1 to 1 with 0 indicating equality) to estimate socioeconomic inequalities in recent (< 12 months) HIV testing uptake [3]. Positivity indicates that HIV testing was more concentrated among the rich while negativity indicates testing was more concentrated among the poor. Inequality estimates were then decomposed using the Recentered Influence Function (RIF) regression method to assess the marginal effect of each country-level factor (i.e., national HIV prevalence and per capita GDP) on the ECI [4]. To do this, we considered the RIF value of the ECI as our dependent variable, which suggests how each individual influences the ECI [4].

We analyzed 16 surveys conducted among 315,847 participants (≥ 15 years old) in Cameroon, Côte d’Ivoire, Congo DR, Ethiopia, Guinea, Kenya, Lesotho, Liberia, Malawi, Mali, Niger, Rwanda, Sierra Leone, Tanzania, Zambia and Zimbabwe. Significantly positive ECI values ranging from 0.03 [95% Confidence Interval 0.01; 0.05] to 0.21 [0.19; 0.23], indicating concentration of HIV testing uptake among the rich, were observed in 12 out of 16 countries. No inequalities were observed in Zimbabwe, Rwanda, Lesotho and Zambia. Figure 1 presents the relationship between country-level ECIs and i) national HIV prevalence; or ii) per-capita GDP. Coefficients from the RIF regression could be interpreted similarly to a standard linear regression. RIF decomposition analysis showed that an increase in HIV prevalence decreased inequality in recent HIV testing (coefficient -5.5 × 10^−3^ 95% CI [-9.6 × 10^−3^; -1.3x 10^−3^]); while GDP per capita had no significant effect on inequality (Coefficient 3.2 ×10^−5^ [-1.1×10^−4^; 4.7×10^−5^]).

**Figure 1.**
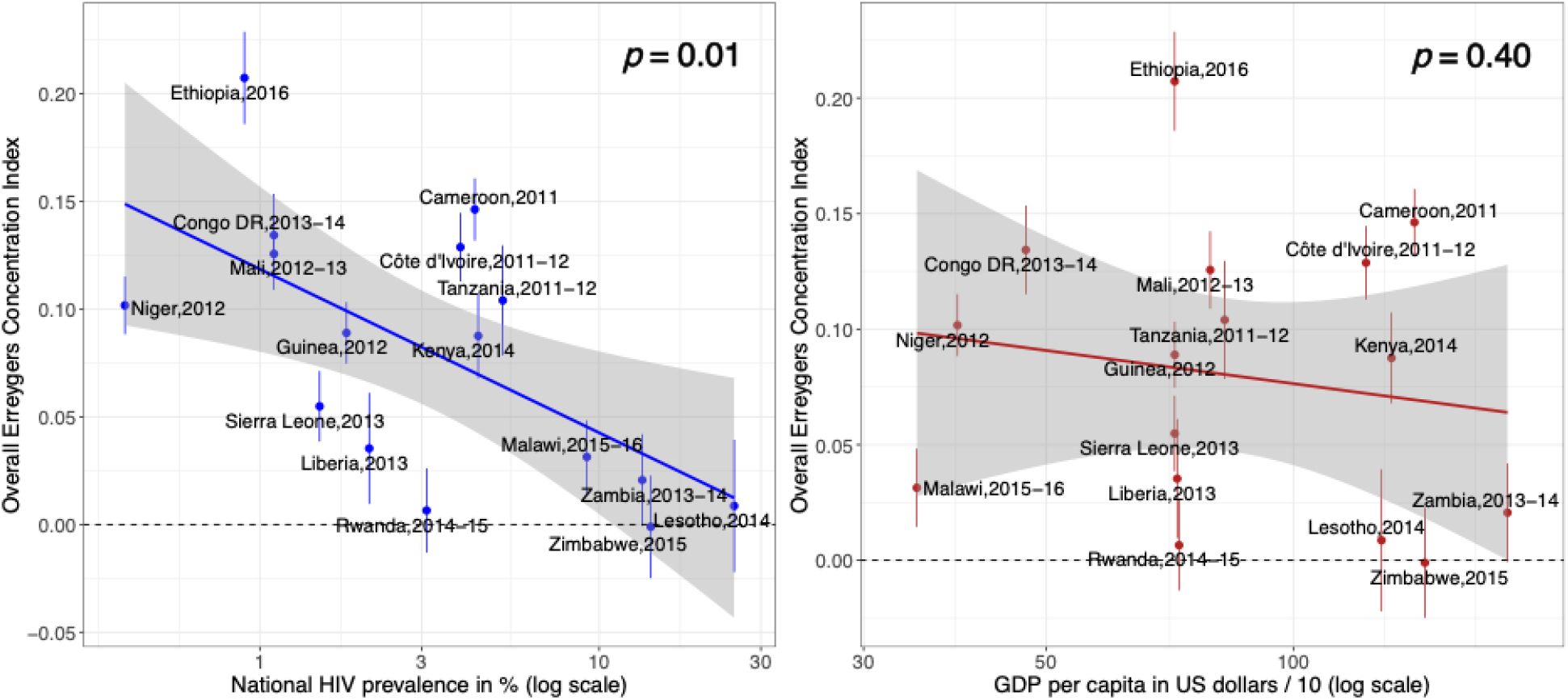
Bivariate linear regression analysis between country-level Erreygers Concentration Index and national HIV prevalence or GDP per capita in US dollars.

Despite the overall increase in HIV testing in recent years, important pro-rich socioeconomic inequalities in recent testing remained in the majority of SSA countries. Our results suggest that the level of the HIV epidemic seems to drive the level of response of HIV testing programs, rather than the GDP per capita of the country (i.e., the national indicator of economic development). These findings are consistent with the patterns we observed in a previous study where countries with low HIV prevalence tended to be the countries with low HIV testing uptake and high pro-rich inequalities [2].

This study has several limitations. We only had 16 country-level estimates in the RIF regression which limited us to only conduct bivariate analysis. Countries also have different survey years, making between-country comparisons not possible. However, our study is among the first to quantify and compare the impact of contextual drivers on inequalities in HIV testing using an innovative methodology and robust data. This novel method had been also utilized in other fields such as in mental health investigating how population changes influenced income-related inequalities in psychiatric diagnoses over time in Sweden [5] and in ageing and health decomposing the effect of factors on health inequality among the elderly in China [6].

In conclusion, our results, which underline the significantly increased socioeconomic inequalities in HIV testing in low prevalence countries, suggest that when responding to the HIV epidemic, there is a need to monitor and assess inequalities in addition to monitoring HIV incidence and prevalence. More research is also needed, integrating a wider range of epidemiological and socioeconomic variables.

## Data Availability

Data from the DHS surveys and World Bank used in this article are publicly available for academic research (www.dhsprogram.com and https://www.worldbank.org/). Processed and formatted data are available upon request from the corresponding author.

https://www.worldbank.org

## Disclaimer

Funding agency had no role in the study design, data collection and analysis.

## References

1. UNAIDS. Fast-Track: Ending the AIDS Epidemic by 2030 [Internet]. 2014. Available from: https://www.unaids.org/sites/default/files/media_asset/JC2686_WAD2014report_en.pdf

2. Ante-Testard PA, Benmarhnia T, Bekelynck A, Baggaley R, Ouattara E, Temime L, et al. Temporal trends in socioeconomic inequalities in HIV testing: an analysis of cross-sectional surveys from 16 sub-Saharan African countries. Lancet Glob Health. 2020 Jun;8(6):e808–18.

3. Erreygers G. Correcting the Concentration Index. J Health Econ. 2009 Mar;28(2):504–15.

4. Heckley G, Gerdtham U-G, Kjellsson G. A general method for decomposing the causes of socioeconomic inequality in health. J Health Econ. 2016 Jul;48:89–106.

5. Linder A, Spika D, Gerdtham U-G, Fritzell S, Heckley G. Education, immigration and rising mental health inequality in Sweden. Soc Sci Med. 2020 Nov;264:113265.

6. Pan, Fan, Yang, Deng. Health Inequality Among the Elderly in Rural China and Influencing Factors: Evidence from the Chinese Longitudinal Healthy Longevity Survey. Int J Environ Res Public Health. 2019 Oct 20;16(20):4018.

